# Risk Factors for New-Onset Atrial Fibrillation in Thai Adults with Hypertension

**DOI:** 10.1101/2025.05.07.25327197

**Authors:** Varisa Limpijankit, Thinnakrit Sasiprapha, Htun Teza, Anuchate Pattanateepapon, Sukanya Siriyotha, Suparee Boonmanunt, John Attia, Ammarin Thakkinstian

**Affiliations:** Chulalongkorn University International Medical Program, The Faculty of Medicine, Chulalongkorn University, Bangkok, Thailand; Division of Cardiology, Department of Medicine, Faculty of Medicine Ramathibodi Hospital, Mahidol University, Bangkok, Thailand; Department of Clinical Epidemiology and Biostatistics, Faculty of Medicine Ramathibodi Hospital, Mahidol University, Bangkok, Thailand; Centre for Clinical Epidemiology and Biostatistics, School of Medicine and Public Health, Faculty of Health and Medicine, University of Newcastle, and Hunter Medical Research Institute, New Lambton, NSW, Australia

**Author notes:** Corresponding author: Thinnakrit Sasiprapha, MD, Division of Cardiology, Department of Medicine, Faculty of Medicine Ramathibodi Hospital, Mahidol University, 270 Rama VI Road, Ratchathewi, Bangkok, Thailand 10400.

**Keywords:** cohort, risk factors, new-onset atrial fibrillation, hypertension, Thailand, statins

## Abstract

**Background:** Limited data exist on new-onset atrial fibrillation (NOAF) risk factors in Asian populations with hypertension (HTN). This study identified predictors of NOAF in Thai adults with HTN.

**Methods:** We conducted a retrospective cohort study of adults (≥18 years) newly diagnosed with HTN at Ramathibodi Hospital, Bangkok (2010–2023). Patients with prior atrial fibrillation or predisposing conditions (e.g., valvular heart disease, hyperthyroidism) were excluded. Baseline demographics, comorbidities, and medication use were analyzed as time-varying covariates using a multivariate Cox proportional hazards model.

**Results:** Of 293,798 HTN patients, 168,441 met inclusion criteria. Over a median 3.7-year follow-up (range: 2.2–8.0), 5,028 developed NOAF (incidence: 5.7 per 1,000 person-years). An age–body mass index (BMI) interaction was observed. In patients <60 years, low BMI increased NOAF risk [hazard ratio (HR) 2.3; 95% CI: 1.4–3.6], while overweight [HR 1.1; 0.8–1.4] and obesity [HR 0.8; 0.6–1.1] showed no significant effect. In patients ≥60 years, NOAF risk rose 2- to 4-fold across BMI categories. Male sex and comorbidities (vascular disease, stroke, heart failure, chronic kidney disease, hyperuricemia) increased risk by 1.2–1.8-fold. Statin use reduced risk [HR 0.8; 0.7–0.9]; sodium-glucose cotransporter-2 inhibitors and glucagon-like peptide-1 receptor agonists showed a non-significant protective trend [HR 0.8; 0.7–1.1].

**Conclusions:** Older age, male sex, abnormal BMI, and the presence of comorbidities are significant risk factors for NOAF in Thai patients with HTN. In contrast, statin use may offer a protective effect.

## Introduction

Systemic arterial hypertension (HTN) is a prevalent chronic non-communicable disease and a leading risk factor for cardiovascular disease and all-cause mortality (1,2). A 2016 World Health Organization (WHO) report estimated that 22% of the global population and 26% of the Southeast Asian population have HTN (3). In Thailand, the 2019 National Health Survey found that 25% of adults have HTN (4), a condition responsible for two-thirds of strokes and half of coronary artery disease events (5).

HTN is a well-established major risk factor for atrial fibrillation (AF) (6,7), contributing to approximately 50% of AF cases (6,8). Long-standing uncontrolled HTN can lead to AF through a complex interplay of factors, including cardiac structural changes (such as left ventricular hypertrophy, left atrial enlargement, and fibrosis), electrical disturbances, neurohormonal activation, and increased sympathetic activity (9,10). Additionally, systemic inflammation, oxidative stress, and endothelial dysfunction are commonly observed in individuals with HTN (11–13). These factors together contribute to the formation of an AF substrate, disrupting the electrical and mechanical functions of the atria and promoting the initiation and maintenance of AF. HTN has been shown to increase the risk of developing AF by 1.8-fold and the risk of progression to permanent AF by 1.5-fold (9).

Both HTN and AF are synergistically associated with an increased incidence of heart failure, stroke, and mortality (10,14). Therefore, identifying predictors of new-onset AF (NOAF) in patients with HTN is essential for preventing or delaying the onset of this arrhythmia. Behavioral modifications could help address these risk factors and combined with guideline-directed medical therapy for HTN, significantly reduce the likelihood of developing AF and its associated complications (15,16).

Several scoring systems have been developed to assess the risk of AF, using community-based cohorts, primarily from Western populations (17–19). However, only one of these risk scores has been specifically validated for patients with HTN: the risk score developed for the ESCARVAL (Estudio CARdiometabolico VALenciano)-RISK study in Spain (20). This score identified age, sex, obesity, and heart failure as independent predictors of NOAF.

AF is a multifactorial condition influenced by aging, genetics, lifestyle, and comorbidities. Identifying independent predictors of AF across diverse populations is crucial for timely intervention and lifestyle modification. However, the specific etiology of NOAF in individuals with HTN, particularly in Asian populations, remains poorly understood (21). Therefore, this study aimed to identify individuals with NOAF in a large cohort of Thai adults with HTN and identify the associated factors, including demographics, comorbidities, and medication use.

## Methods

### Study design & subjects

This retrospective cohort analysis utilized data from the Clinical Epidemiology and Biostatistics (CEB) HTN Data Warehouse at the Faculty of Medicine Ramathibodi Hospital, Mahidol University (Bangkok, Thailand). The database included electronic medical records of routine patients from January 2010 to December 2023. Our analysis focused on demographic information, comorbidities, and medication usage. The study protocol was approved by the Human Research Ethics Committee of the Faculty of Medicine Ramathibodi Hospital (COA.MURA 2024/681).

The CEB database for 2010-23 contained 293,798 patients with HTN diagnosed at Ramathibodi Hospital. These patients were initially identified based on ICD-10 codes and/or the use of anti-hypertensive medications, such as angiotensin-converting enzyme inhibitors (ACEIs), angiotensin receptor blockers (ARBs), calcium channel blockers (CCBs), beta-blockers (BBs), alpha-blockers, and diuretics. Subjects included in our analysis met the following inclusion criteria: (1) age 18 years or older; (2) newly diagnosed with HTN without any complications; (3) no prior medical history of AF; and (4) at least one follow-up visit more than 30 days post-diagnosis. The exclusion criterion was the presence of pre-existing conditions that could predispose individuals to AF, such as valvular heart disease or hyperthyroidism.

### Data Collection

Covariates potentially associated with NOAF were extracted from the master cohort database, including age, sex, body mass index (BMI, kg/m^2^), and comorbidities at baseline and follow-up (using ICD-10 codes, as detailed in Supplementary Table 1). The following definitions were used: diabetes mellitus (DM), defined by fasting plasma glucose ≥ 126 mg/dL, HbA1c ≥6.5% on two consecutive tests, or the use of anti-diabetic medications (22); dyslipidemia, defined by total cholesterol ≥ 200 mg/dL, LDL-C ≥ 130 mg/dL, or the use of statins or other lipid-lowering medications; chronic kidney disease (CKD), defined by an estimated glomerular filtration rate (eGFR) < 60 mL/min/1.73 m^2^ on two consecutive occasions within 90 days; a history of vascular diseases (including coronary artery disease, upper and lower extremity peripheral arterial disease, carotid disease, renal artery disease, and aortic disease; heart failure, determined by echocardiogram with a left ventricular ejection fraction <40% or use of guideline-directed medical therapy; stroke (23); obstructive sleep apnea (OSA); and hyperuricemia, defined as elevated serum uric acid levels > 6 mg/dL in women and > 7 mg/dL in men, or the use of uric-lowering medications. These comorbidities were cross-validated with other fields from the CEB Warehouse data set, where available.

Medication usage data were retrieved from prescriptions and pharmacy dispensing records. These included antihypertensive drugs (as listed above), mineralocorticoid receptor antagonists (MRAs, e.g., spironolactone), diabetic medications [e.g., sulfonylureas, metformin, thiazolidinediones, dipeptidyl peptidase-4 inhibitors, alpha-glucosidase inhibitors, sodium-glucose cotransporter-2 inhibitors (SGLT2i), glucagon-like peptide-1 receptor agonist (GLP-1RA), and insulin], statins, antiarrhythmic drugs (e.g., amiodarone, digoxin, non-dihydropyridine CCBs such as verapamil and diltiazem), antiplatelet agents (e.g., aspirin, clopidogrel, ticagrelor, prasugrel), and oral anticoagulants (OACs) (e.g., warfarin and direct oral anticoagulants). All medications were classified by their generic names and drug classes.

Data on these variables, collected at baseline and during follow-up visits prior to the development of NOAF (treated as time-varying covariates), were included in the analysis.

### Outcomes of Interest and Follow-Up

NOAF was the primary outcome of this analysis. It was identified using the CEB AF data warehouse (Supplementary Figure 1) through the following steps: 1) data were retrieved from three sources: ICD-10 codes (I48*), ECG/Holter reports, and prescriptions of OACs (warfarin, dabigatran, rivaroxaban, apixaban, edoxaban); 2) ICD-10 and ECG/Holter data were linked using hashed hospital number and date (within 90 days); 3) patients with ECG/Holter-confirmed AF (with or without ICD-10 codes) were considered NOAF cases; 4) patients diagnosed with ICD-10 alone were linked to OAC prescriptions and any OAC use was confirmed in NOAF patients. Those without OAC use were verified via electrical medical records. The follow-up end date for each patient was either the NOAF diagnosis date (ICD-10 or ECG/Holter, whichever came first) or December 31, 2023. Time to NOAF or censoring was then calculated by subtracting the end date from the baseline date (the date when HTN was diagnosed).

### Statistical analysis

Continuous data were expressed as means with standard deviations (SDs), while categorical data were presented as counts (n) and percentages. A Cox proportional hazards model with time-varying covariates was used to identify factors associated with NOAF. Initially, a univariate Cox model was constructed by fitting each covariate to NOAF. Covariates with p-values less than 0.20 were then included in a multivariate Cox model. A likelihood ratio test was used to select and retain only the significant covariates for the final model. Potential interaction effects between covariates were also explored where appropriate. The probabilities of NOAF occurrence were estimated, and hazard ratios (HRs) with 95% confidence intervals (CIs) were calculated. A sensitivity analysis was subsequently performed, considering death from any cause as a competing risk event. The Fine and Gray sub-hazard model (SHD) with time-varying covariates was applied to minimize survival bias related to the occurrence of NOAF, accounting for the possibility that individuals may have died before developing NOAF. Sub-distribution hazard ratios (SHRs) and 95% CIs were estimated. All statistical analyses were conducted using RStudio and STATA 18, with a p-value of < 0.05 considered statistically significant.

## Results

### Baseline characteristics of subjects

Of the 293,798 patients initially identified with HTN (Supplementary Figure 2), 168,441 met the inclusion criteria and were included in the cohort for analysis (Figure 1). The baseline characteristics of 63,045 newly diagnosed hypertensive patients were available and are presented in Table 1. The cohort was predominantly female (58.7%), with a mean age of 59.5±14.6 years. Most patients were aged 60–79 years (48.1%) or over 80 years (8.2%). Based on the BMI cutoffs for Asians, 36.8% of participants were classified as overweight (BMI > 23 kg/m²), and 33.4% were classified as obese (BMI > 27 kg/m²). The prevalences of comorbid conditions were as follows: type 2 DM (20.4%), dyslipidemia (4.5%), CKD (13.2%), vascular diseases (15.0%), heart failure (0.2%), stroke (4.2%), and hyperuricemia (13.1%).

**Figure 1.**
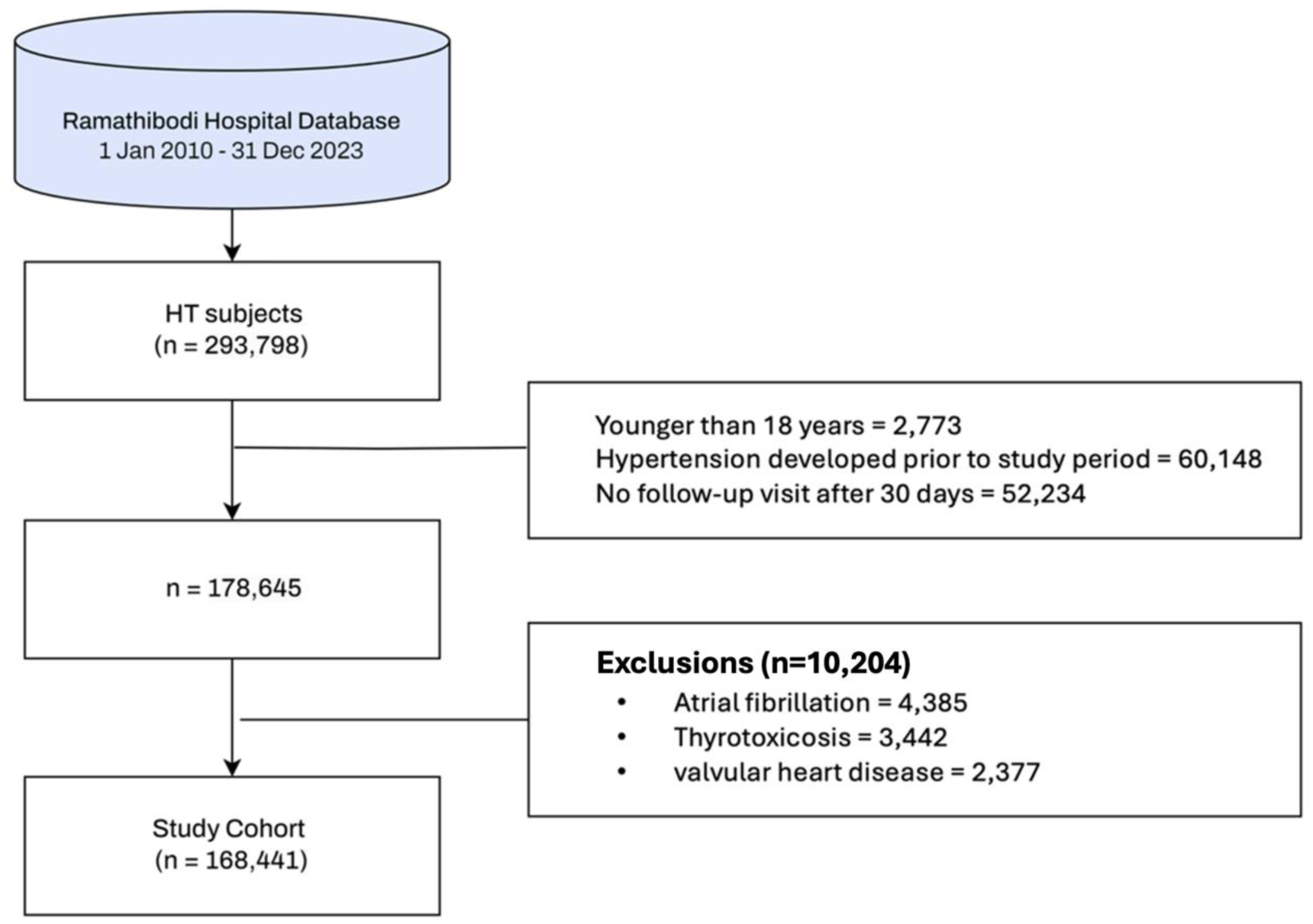
Study Flowchart.

**Table 1.** Distribution of Baseline and Time-Varying Data.

| Features | Baseline Data<br>(n = 168,441) | Time-varying Data<br>(N = 564,644) |
| --- | --- | --- |
| Age, yrs, (mean±SD) | 59.5±14.6 | 61.8±14.2 |
| < 60, (%) | 43.7 | 37.4 |
| 60 – 79, (%) | 48.1 | 52.4 |
| ≥ 80, (%) | 8.2 | 10.2 |
| Female, (%) | 58.7 | 58.7 |
| Male, (%) | 41.3 | 41.3 |
| BMI, kg/m<sup>2</sup>, (mean±SD) | 25.6±4.8 | 26.2± 4.9 |
| < 18.5, (%) | 4.1 | 3.2 |
| 18.5 – 22.9, (%) | 25.7 | 22.4 |
| 23.0 – 26.9, (%) | 36.8 | 36.3 |
| ≥ 27, (%) | 33.4 | 38.2 |
| **Comorbidities** |  |  |
| Vascular diseases, (%) | 15.0 | 26.3 |
| Stroke, (%) | 4.2 | 5.9 |
| Heart failure, (%) | 0.2 | 1.0 |
| Dyslipidemia, (%) | 4.5 | 8.4 |
| CKD, (%) | 13.2 | 13.2 |
| OSA, (%) | 4.0 | 6.7 |
| Hyperuricemia, (%) | 13.1 | 13.4 |
| **Medications** |  |  |
| Antihypertensive (%) |  |  |
| None | 38.7 | 46.0 |
| Monotherapy | 37.0 | 30.3 |
| Combination therapy | 24.3 | 23.7 |
| Type 2 DM (%) |  |  |
| No | 79.6 | 72.9 |
| Yes, with other medications | 19.2 | 24.9 |
| Yes, with SGLT2i or GLP-1 agonist | 1.2 | 2.3 |
| MRAs, (%) | 1.8 | 1.8 |
| Statins, (%) | 36.9 | 42.8 |
| Antiarrhythmic agents, (%) | 2.2 | 1.7 |
| Anticoagulants, (%) | 4.7 | 3.3 |
| Antiplatelets, (%) | 13.7 | 13.4 |

n = Number of patients; N = Number of observations
Abbreviation: BMI, body mass index; CKD, chronic kidney disease; DM, diabetes mellitus; GLP, glucagon-like peptide; MRAs, mineralocorticoid receptor antagonists; OSA, obstructive sleep apnea; SGLT2i, sodium-glucose Cotransporter-2 inhibitor.

At baseline, most patients (61.3%) were treated with antihypertensive medications, with 37.0% receiving monotherapy and 24.3% receiving combination therapy. In contrast, 38.7% of patients were recommended lifestyle modifications alone, without antihypertensive drugs. MRAs were used by only 1.8% of patients, and one-third of patients were treated with statins. All diabetic patients were on medications, with 1.2% receiving SGLT2i or GLP-1RA. Antithrombotic agents were prescribed to 18.4% of patients (13.7% received antiplatelets, and 4.7% received OACs), while 2.2% of patients were using antiarrhythmic drugs.

Data from 99,408 patients were available during the follow-up period. When comparing baseline and time-varying data (Table 1), there were increases in the frequencies of older age, overweight/obesity, and comorbidities, including vascular disease (from 15.0% to 25.3%), stroke (from 4.2% to 5.9%), heart failure (from 0.2% to 1.0%), OSA (from 4.0% to 6.7%), and type 2 DM (from 19.1% to 27.2%). Regarding treatments during follow-up, the use of statins and antidiabetic medications increased over time, but there were no significant changes in the use of MRAs, antihypertensive drugs, antithrombotic agents, or antiarrhythmic drugs.

### Incidence and Predictors of NOAF

Of 168,441 patients with HTNs, 5,028 (3.0%) were diagnosed with NOAF during follow-up (median of 3.7 years; range 2.2–8.0), yielding an incidence of 5.7 per 1,000 person-years. Univariate analysis for time-varying covariates (n=99,408) identified age as one of the strongest risk factors for the occurrence of NOAF (Table 2). Patients aged ≥80 years had a hazard ratio (HR) of 6.4 (95% CI: 5.6–7.3), while those aged 60-79 years had an HR of 2.4 (2.2–2.7). Male gender was also associated with increased risk [HR 1.4 (1.3–1.5)]. Unexpectedly, low body weight (BMI <18.5 kg/m^2^) was linked to higher NOAF risk [HR 1.5 (1.2-1.8], while overweight and obesity were associated with lower risk (HRs ∼0.9 vs. normal BMI).

**Table 2.** Univariate Analysis of Time-varying Covariates.

| Factors | No AF<br>(N=163,415) | AF<br>(N=5,026) | Total time-<br>at-risk | Incidence Rate per<br>1,000 person-years | Hazard Ratios |
| --- | --- | --- | --- | --- | --- |
| Age, yrs, (mean±SD) | 59.5±14.6 | 68.3±13.3 |  |  |  |
| < 60 | 72,554 | 1,086 | 426806.6 | 2.5 (2.4, 2.7) | 1 |
| 60 – 79 | 78,175 | 2,840 | 391592.4 | 7.2 (7.0, 7.5) | 2.4 (2.2 – 2.7) |
| ≥ 80 | 12,686 | 1,100 | 55825.8 | 19.7 (18.6, 20.9) | 6.4 (5.6 – 7.3) |
| Female | 96,392 | 2,410 | 524658.8 | 4.6 (4.4, 4.8) | 1 |
| Male | 67,023 | 2,616 | 349566.1 | 7.5 (7.2, 7.8) | 1.4 (1.3 – 1.5) |
| BMI, kg/m<sup>2</sup>, (mean±SD) | 25.6±4.8 | 24.6±4.6 |  |  |  |
| < 18.5 | 4535 | 208 | 19050.31 | 10.92 (9.48, 12.51) | 1.5 (1.2 – 1.8) |
| 18.5 – 22.9 | 28875 | 907 | 117698.22 | 7.71 (7.21, 8.22) | 1 |
| 23.0 – 26.9 | 41476 | 1105 | 164563.62 | 6.71 (6.32, 7.12) | 0.9 (0.8 – 1.0) |
| ≥ 27 | 37844 | 793 | 146723.04 | 5.40 (5.04, 5.79) | 0.9 (0.8 – 1.0) |
| **Comorbidities** |  |  |  |  |  |
| Vascular diseases, (%) |  |  |  |  |  |
| No | 139,512 | 3,641 | 772553.2 | 4.7 (4.6, 4.9) | 1 |
| Yes | 23,903 | 1,385 | 101671.7 | 13.6 (12.9, 14.4) | 2.5 (2.3 – 2.7) |
| Stroke, (%) |  |  |  |  |  |
| No | 156,775 | 4,629 | 846923.2 | 5.5 (5.3, 5.6) | 1 |
| Yes | 6,640 | 397 | 27301.7 | 14.5 (13.1, 16.0) | 1.9 (1.7 – 2.2) |
| Heart Failure, (%) |  |  |  |  |  |
| No | 163,157 | 4,988 | 873594.2 | 5.7 (5.5, 5.9) | 1 |
| Yes | 258 | 38 | 630.7 | 60.2 (42.6, 82.7) | 5.2 (4.3 – 6.4) |
| CKD, (%) |  |  |  |  |  |
| No | 151,964 | 4,179 | 832365.7 | 5.0 (4.9, 5.2) | 1 |
| Yes | 11,451 | 847 | 41859.2 | 20.2 (18.9, 21.6) | 1.0 (0.8 – 1.1) |
| OSA, (%) |  |  |  |  |  |
| No | 156,742 | 4,891 | 850108.6 | 5.7 (5.6, 5.9) | 1 |
| Yes | 6,673 | 135 | 24116.3 | 5.6 (4.7, 6.6) | 3.2 (2.9 – 3.5) |
| Hyperuricemia, (%) |  |  |  |  |  |
| No | 142,401 | 3,947 | 776919.7 | 5.1 (4.9, 5.2) | 1 |
| Yes | 21,014 | 1,079 | 97305.2 | 11.1 (10.4, 11.8) | 1.9 (1.7 – 2.1) |
| **Medications** |  |  |  |  |  |
| Antihypertensive, (%) |  |  |  |  |  |
| None | 64,126 | 1,127 | 445885.0 | 2.5 (2.4, 2.7) | 1 |
| Monotherapy | 60,768 | 1,633 | 277052.9 | 5.9 (5.6, 6.2) | 1.8 (1.6 – 2.0) |
| Combination therapy | 38,521 | 2,266 | 151287.0 | 15.0 (14.4, 15.6) | 3.3 (3.0 – 3.6) |
| Type 2 DM, (%) |  |  |  |  |  |
| No | 130,374 | 3,791 | 727897.7 | 5.2 (5.0, 5.4) | 1 |
| Yes, with other medications | 31,036 | 1,169 | 141272.0 | 8.3 (7.8, 8.8) | 1.2 (1.1 – 1.3) |
| Yes, with SGLT2i or GLP-1 agonist | 2,005 | 66 | 5055.2 | 13.1 (10.1, 16.6) | 1.4 (1.1 – 1.7) |
| MRAs, (%) |  |  |  |  |  |
| No | 160,824 | 4,633 | 862888.2 | 5.4 (5.2, 5.5) | 1 |
| Yes | 2,591 | 393 | 11336.7 | 34.7 (31.3, 38.3) | 3.6 (3.0 – 4.3) |
| Statins, (%) |  |  |  |  |  |
| No | 103,768 | 2,493 | 613140.7 | 4.1 (3.9, 4.2) | 1 |
| Yes | 59,647 | 2,533 | 261084.2 | 9.7 (9.3, 10.1) | 1.5 (1.4 – 1.6) |
| Antiarrhythmic agents, (%) |  |  |  |  |  |
| No | 160671 | 4010 | 860013.8 | 4.67 (4.53, 4.82) | 1 |
| Yes | 2744 | 1006 | 14211.12 | 70.79 (66.48, 75.30) | 6.5 (5.6 – 7.6) |
| Anticoagulants, (%) |  |  |  |  |  |
| No | 157038 | 3422 | 841556.74 | 4.07 (3.93, 4.20) | 1 |
| Yes | 6377 | 1604 | 32668.17 | 49.10 (46.73, 51.56) | 9.8 (8.8 – 10.8) |
| Antiplatelets, (%) |  |  |  |  |  |
| No | 141988 | 3347 | 763054.29 | 4.39 (4.24, 4.54) | 1 |
| Yes | 21427 | 1679 | 111170.63 | 15.10 (14.39, 15.84) | 2.3 (2.1 – 2.6) |

Abbreviation as in Table 1

Comorbidities associated with higher NOAF risk included pre-existing vascular diseases [HR 2.5 (2.3–2.7)], stroke [HR 1.9 (1.7–2.2)], heart failure [HR 5.2 (4.3–6.4)], CKD [HR 1.0 (4.3–6.4)], OSA [HR 3.2 (2.9–3.5)], and hyperuricemia [HR 1.9 (1.7–2.1)].

Regarding treatments, combination antihypertensive therapy was more strongly associated with NOAF than monotherapy (both compared to no treatment), with HRs of 3.3 (3.0–3.6) and 1.8 (1.6–2.0), respectively. Other medications associated with NOAF included MRAs [HR 3.6 (3.0–4.3)], antidiabetic medications [HR 1.2 (1.1–1.3)], antidiabetic medications combined with SGLT2i or GLP-1RA [HR 1.4 (1.1–1.7)], and statins [HR 1.5 (1.4–1.6)]. All antithrombotic and antiarrhythmic agents were also associated with a higher risk of NOAF, as shown in Table 2.

A multivariate time-varying Cox model (Table 3 and Figure 2) identified 11 factors significantly associated with increased NOAF risk: advanced age, male sex, BMI, five pre-existing comorbidities (vascular disease, heart failure, CKD, hyperuricemia, and stroke), antihypertensive medication use, type 2 diabetes, and statin use.

**Figure 2.**
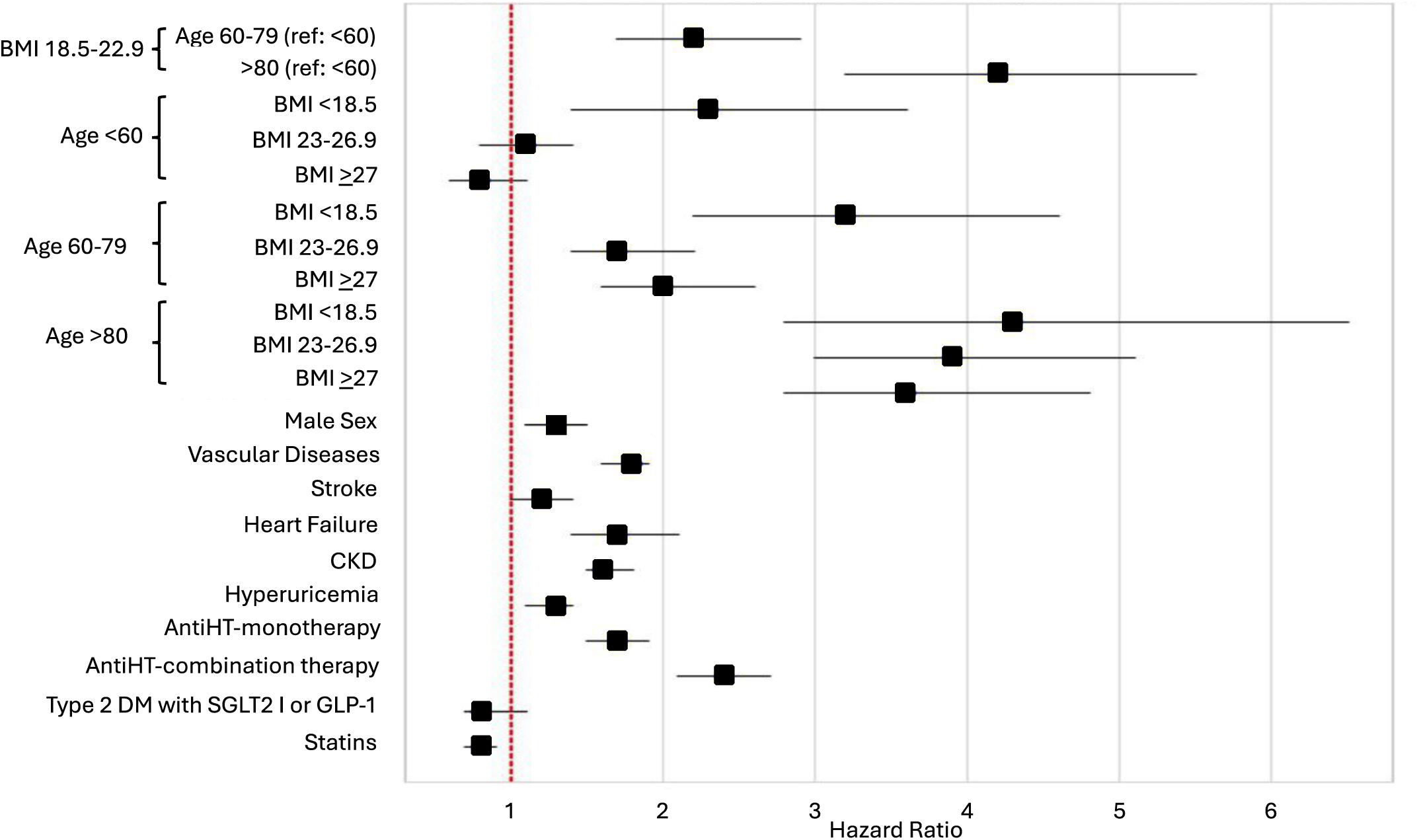
Forest Plot of factors associated with AF in hypertension: A multivariable time-varying Cox model.

**Table 3.**
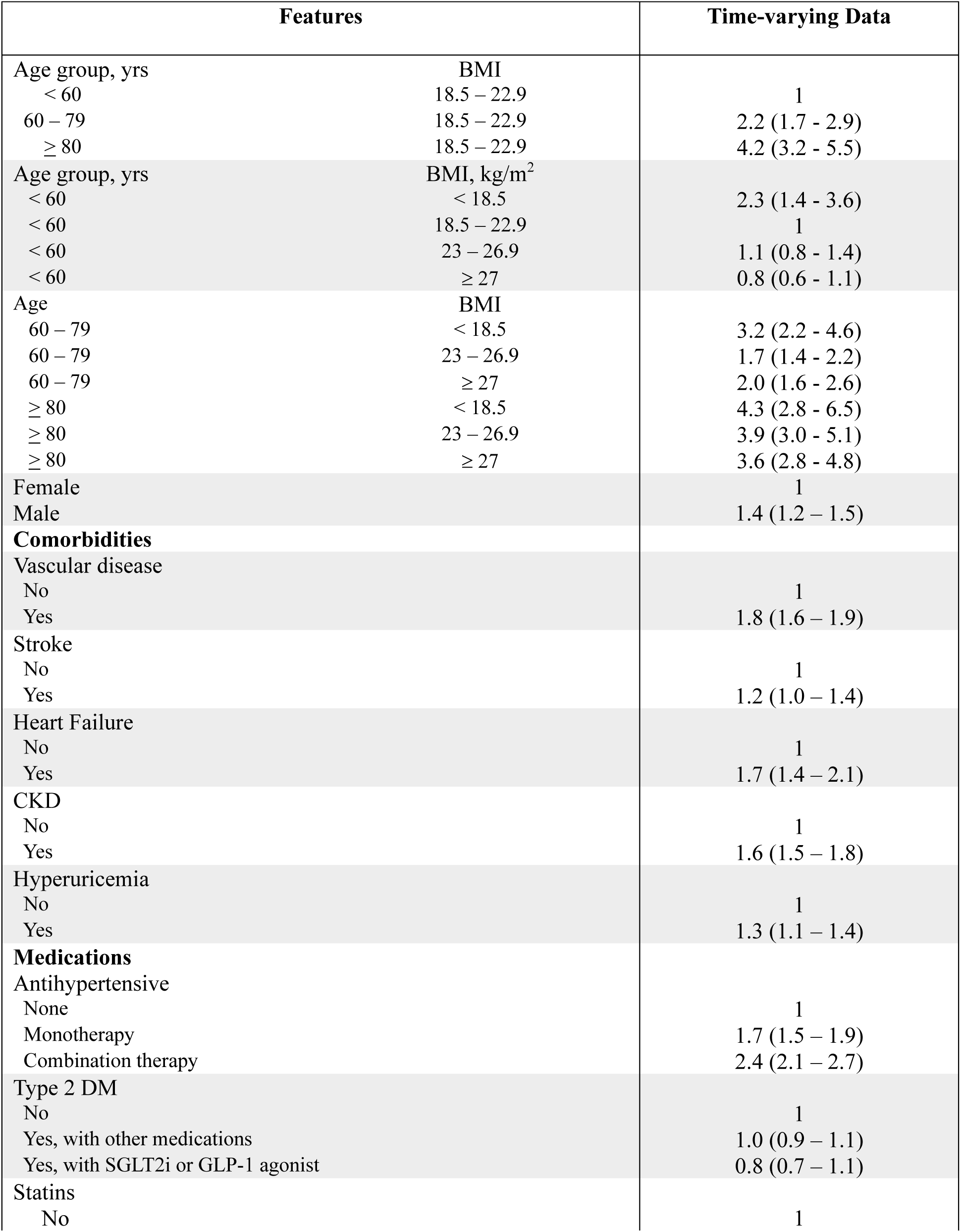
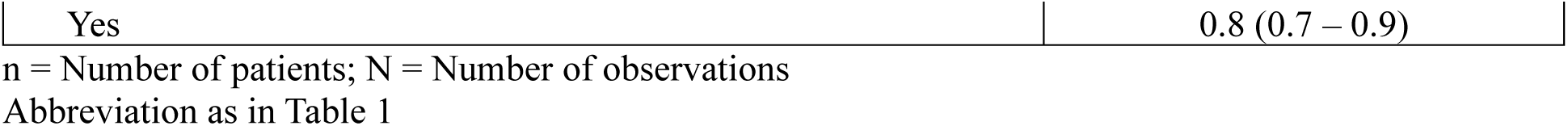
Factors associated with AF using time-varying data: A Multivariate Cox Proportional Hazards Model.

| Features |  | Time-varying Data |
| --- | --- | --- |
| Age group, yrs | BMI |  |
| < 60 | 18.5 – 22.9 | 1 |
| 60 – 79 | 18.5 – 22.9 | 2.2 (1.7 - 2.9) |
| ≥ 80 | 18.5 – 22.9 | 4.2 (3.2 - 5.5) |
| Age group, yrs | BMI, kg/m<sup>2</sup> |  |
| < 60 | < 18.5 | 2.3 (1.4 - 3.6) |
| < 60 | 18.5 – 22.9 | 1 |
| < 60 | 23 – 26.9 | 1.1 (0.8 - 1.4) |
| < 60 | ≥ 27 | 0.8 (0.6 - 1.1) |
| Age | BMI |  |
| 60 – 79 | < 18.5 | 3.2 (2.2 - 4.6) |
| 60 – 79 | 23 – 26.9 | 1.7 (1.4 - 2.2) |
| 60 – 79 | ≥ 27 | 2.0 (1.6 - 2.6) |
| ≥ 80 | < 18.5 | 4.3 (2.8 - 6.5) |
| ≥ 80 | 23 – 26.9 | 3.9 (3.0 - 5.1) |
| ≥ 80 | ≥ 27 | 3.6 (2.8 - 4.8) |
| Female |  | 1 |
| Male |  | 1.4 (1.2 – 1.5) |
| **Comorbidities** |  |  |
| Vascular disease |  |  |
| No |  | 1 |
| Yes |  | 1.8 (1.6 – 1.9) |
| Stroke |  |  |
| No |  | 1 |
| Yes |  | 1.2 (1.0 – 1.4) |
| Heart Failure |  |  |
| No |  | 1 |
| Yes |  | 1.7 (1.4 – 2.1) |
| CKD |  |  |
| No |  | 1 |
| Yes |  | 1.6 (1.5 – 1.8) |
| Hyperuricemia |  |  |
| No |  | 1 |
| Yes |  | 1.3 (1.1 – 1.4) |
| **Medications** |  |  |
| Antihypertensive |  |  |
| None |  | 1 |
| Monotherapy |  | 1.7 (1.5 – 1.9) |
| Combination therapy |  | 2.4 (2.1 – 2.7) |
| Type 2 DM |  |  |
| No |  | 1 |
| Yes, with other medications |  | 1.0 (0.9 – 1.1) |
| Yes, with SGLT2i or GLP-1 agonist |  | 0.8 (0.7 – 1.1) |
| Statins |  |  |
| No |  | 1 |

|  |  |
| --- | --- |
| Yes | 0.8 (0.7 – 0.9) |

n = Number of patients; N = Number of observations
Abbreviation as in Table 1

An interaction effect between age and BMI was observed (Supplementary Table 2). The mean age (±SD) was lower among obese patients (60.4 ± 13.3 years) and higher among those with low BMI (64.1 ± 18.9 years) compared to patients with normal BMI (62.5 ± 15.5 years). Additionally, the proportion of patients under 60 years of age was highest in the obese group (41.5%), while the proportion of patients over 80 years of age was highest in the low BMI group (24.2%). Based on these findings, an interaction term between age and BMI was included in the final model (Likelihood ratio: Chi-square = 17.1 (df=6), p = 0.009).

Among patients with normal BMI, advanced age was the strongest predictor of NOAF, followed by vascular disease, CHF, CKD, hyperuricemia, and stroke. In individuals under 60 with normal BMI, low BMI was significantly associated with increased NOAF risk [HR 2.3 (1.4–3.6)], while overweight showed a non-significant slight increase in risk [HR 1.1 (0.8–1.4)]. In contrast, obesity was associated with a non-significant trend toward reduced risk [HR 0.8 (0.6–1.1)].

In patients aged 60 years or older, the risk of NOAF increased significantly by approximately 2- to 3-fold among those aged 60–79 with low BMI or who were overweight or obese and by about 3- to 4-fold in patients aged 80 and above across all three BMI categories, compared to those with normal BMI.

The presence of comorbidities (vascular disease, CHF, CKD, hyperuricemia, or stroke) was associated with a 1.2- to 1.8-fold increased risk of NOAF. Antihypertensive monotherapy and combination therapy were linked to 1.7-fold and 2.4-fold increased risks, respectively. Conversely, statin use was associated with a 20% reduction in NOAF risk [HR 0.8 (0.7–0.9)]. Combined antidiabetic therapy involving SGLT2i or GLP-1RA showed a non-significant trend toward reduced risk [HR 0.8 (0.7–1.1)].

A sensitivity analysis using a competing risk model was conducted. The overall death rate was 1.5 per 1000 patients per year. The SHRs and their corresponding 95% CIs for risk factors were estimated and showed similar effect sizes to those found in the survival model (see Supplementary Table 3).

## Discussion

This large-scale, real-world cohort study of 168,441 hypertensive patients in Thailand, with a median follow-up of 3.7 years, provides critical insights into the incidence and risk factors of NOAF. Our findings identified eight factors associated with increased risk of NOAF, some aligning with current literature, while others revealed novel or unexpected associations. This study provides valuable insights into the specific risk factors for NOAF in Thai and Southeast Asian populations, offering crucial data to guide clinical management for these at-risk groups. Notably, statin use was linked to a reduced risk of NOAF, while the effects of SGLT2i and GLP-1RA were not statistically significant.

The overall incidence of NOAF in this cohort was 5.7 per 1,000 person-years, which is lower than published global estimates (6.3-9.9 per 1,000 person-years) (17,24,25). This discrepancy may be attributed to factors such as differences in healthcare access, lifestyle, genetic background, HTN management in Thailand, and the relatively short median follow-up period (3.7 years) compared to other studies (7-10 years). Shorter follow-up periods may not capture all NOAF cases, as some cases develop over a longer duration. Despite the lower incidence, NOAF remains a significant concern due to its impact on long-term cardiovascular outcomes, underscoring the need for longer-term studies to better understand lifetime risks.

Consistent with previous studies, older age and male gender were identified as significant risk factors for the development of NOAF in hypertensive patients (17,20,21,26). In older individuals, the mechanisms underlying AF primarily involve structural changes in the atria due to aging, such as atrial fibrosis, reduced cellular connectivity, loss of atrial muscle mass, and electrical remodeling (27). These changes create a substrate that promotes irregular heart rhythms, often exacerbated by altered autonomic function and comorbidities that are more common in older individuals and men. Both electrical remodeling and the effect of sex hormones, along with structural changes like atrial fibrosis, contribute to gender differences in AF (28). Further research—including the inclusion of smoking and alcohol use— is needed to fully understand the mechanisms behind these gender-based disparities in AF development. Our findings highlight the importance of early detection and management of AF risk factors in older hypertensive male patients, especially those with multiple comorbidities, to reduce the risk of NOAF and its associated complications.

Another important finding in our study was the association between pre-existing comorbidities (i.e., vascular diseases, heart failure, and stroke) and an increased risk of NOAF. These comorbid conditions likely contribute to the development of AF through well-established pathophysiologic mechanisms, including increased left atrial pressure, atrial fibrosis, and altered blood flow dynamics. These risk factors are integral components of the CHA2DS2-VASc score, which is used to predict the risk of stroke in AF patients (29). The CHA2DS2-VASc score has also been directly associated with the incidence of NOAF in the general population. Elderly patients with significant changes in their CHA2DS2-VASc score are more likely to develop incident AF (30). These findings emphasize the importance of careful monitoring and comprehensive management of hypertensive patients, addressing not only BP but also these underlying comorbidities to mitigate the risk of NOAF.

Similar to previous studies (31,32), we found that CKD and hyperuricemia are additional risk factors for NOAF in hypertensive patients. CKD activates the renin-angiotensin-aldosterone and sympathetic nervous systems while also inducing oxidative stress, systemic inflammation, and volume overload (33). CKD can lead to hyperuricemia by reducing uric acid excretion, and both conditions often share common underlying pathologies. Hyperuricemia, in turn, contributes to oxidative stress, inflammation, atrial fibrosis, and potential disruption of electrical signaling (34). These factors strongly correlate with atrial electrical changes and structural remodeling, which can promote and sustain AF. The possible role of uric acid-lowering agents as an upstream therapy for HTN to reduce the burden of AF is an important issue for future research.

Effective control of HTN is crucial for reducing the risk of AF. However, we observed an increased risk of NOAF in hypertensive patients using multiple antihypertensive medications. This may be due to factors such as the severity of HTN, comorbidities, clinical inertia, lack of adherence, electrolyte imbalances, autonomic regulation, and/or drug interactions. Some antihypertensive medications, such as diuretics and alpha-blockers, may raise AF risk by affecting electrolyte balance (35). Alpha-blockers, in particular, can cause orthostatic hypotension, potentially triggering arrhythmias (36). In contrast, ACEIs, ARBs, and BBs may offer protection against AF (37). The beneficial effects of ACEIs and ARBs on the development of AF are thought to be related to atrial electrical remodeling, while BBs help lower BP and reduce heart rate, especially in patients with HTN and rapid AF. Further research is needed to evaluate the benefits of different antihypertensive therapies in preventing AF.

The Systolic Blood Pressure Intervention Trial (SPRINT) (38) demonstrated that lowering systolic BP to a target of less than 120 mm Hg (intensive BP lowering) significantly reduced the incidence of cardiovascular events, including AF, compared to a standard target of less than 140 mm Hg in hypertensive patients. This underscores the benefit of more aggressive BP control in reducing cardiovascular events, including AF. In our study, the average number of antihypertensive medications remained unchanged from baseline to follow-up, possibly reflecting a lack of dosage adjustment. Therefore, careful management of HTN—including determining the optimal BP target, selecting appropriate medications, adjusting dosages, and monitoring for adverse effects —particularly in the elderly, may be crucial to minimizing the risk of AF.

Our study found that statin use was associated with a reduced risk of NOAF, aligning with previous research that links statin therapy to a lower incidence of both the onset and recurrence of AF (39,40). In addition to their well-known cholesterol-lowering effects, statins possess pleiotropic properties—including anti-inflammatory, anti-fibrotic, and endothelial-protective actions—which may help prevent atrial remodeling and reduce the risk of arrhythmias (41). These findings suggest that statins may play a role in preventing NOAF in hypertensive patients, offering benefits beyond their traditional cardiovascular effects. However, the evidence remains inconclusive, particularly among individuals without ischemic vascular disease (42). Although longitudinal studies in population cohorts support the role of statins in the primary prevention of AF (43,44), their effect on reducing NOAF specifically in hypertensive populations remains uncertain. Further randomized controlled trials (RCTs) are needed to clarify this relationship. Nonetheless, statins should be considered for patients who develop NOAF, as their use has been associated with a lower risk of ischemic and hemorrhagic stroke, transient ischemic attack, all-cause mortality, and major cardiovascular events (45).

SGLT2i or GLP-1RA, known for their glucose-lowering and cardiovascular protective effects (46–48), were associated with a reduced risk of NOAF in our study. However, the results were not statistically significant. Both drug classes improve endothelial function, reduce inflammation, and help attenuate heart failure progression—mechanisms that may contribute to a lower incidence of AF (49,50). The lack of statistical significance may be due to these drugs’ novelty and high cost, resulting in low usage (1.2%) and a limited sample size, thus insufficient power to detect protective effects. Recent meta-analyses of RCTs have shown that SGLT2i or GLP-1RA significantly reduces the risk of NOAF in both diabetic and non-diabetic patients (51–53). However, the potential benefit of these drugs in reducing AF risk in hypertensive patients requires further investigation in cohorts with longer periods of follow-up.

A novel and clinically relevant finding was the age-dependent association between BMI and NOAF. Among individuals under 60, low BMI was linked to higher NOAF risk. Conversely, in older adults, overweight and obesity were associated with increased NOAF risk, consistent with evidence linking adiposity to atrial enlargement, diastolic dysfunction, and pro-inflammatory states (20,54,55). These results underscore the need for age-specific risk stratification, as uniform BMI thresholds may be insufficient for accurate cardiovascular risk prediction.

Both underweight and obese individuals are at increased risk of AF, with underweight status emerging as an independent and previously under-recognized risk factor. This finding is supported by a large-scale, nationwide study conducted in Korea, which analyzed health data from 132,063 individuals aged 40 and older over a median follow-up period of 9 years (56). The study found a U-shaped relationship between BMI and AF risk: each 1.0 kg/m^2^ decrease below a BMI of 20 increased the risk of AF by 13%, while each 1.0 kg/m^2^ increase above 20 raised the risk by 6%. Compared to individuals of normal weight, those who were underweight had a 23% higher risk of developing AF. The elevated risk in underweight individuals may be associated with factors such as frailty, malnutrition, or chronic illness—conditions that can contribute to autonomic dysregulation and atrial remodeling. Furthermore, a sensitivity analysis using a competing risk model, with death as a competing event, showed similar trends in age-BMI effects, suggesting that survival bias due to death before AF onset did not influence the results. Additional factors, such as thyroid disease, chronic lung disease, cancer, smoking, alcohol use, and physical activity, may also play a role. These findings highlight the importance of maintaining a healthy BMI for the prevention of AF (57).

Another important but potentially underrecognized factor is OSA, which contributes to AF through intermittent hypoxia and sympathetic activation (58,59). While OSA was not significantly associated with NOAF in our study, it may have been underdiagnosed. Prior evidence shows that continuous positive airway pressure therapy can reduce AF recurrence by 42% (60), underscoring the need for better screening and management of OSA in hypertensive patients.

This study fills a crucial knowledge gap by providing region-specific data on NOAF in Asian hypertensive populations. It supports a comprehensive, individualized approach to AF prevention, emphasizing optimal hypertension control, weight management, treatment of comorbidities, and appropriate use of cardiovascular medications.

### Study Limitations

This is the first large cohort study to investigate factors associated with atrial fibrillation (AF) in Asian patients with hypertension. However, several limitations should be acknowledged. First, as a retrospective cohort study conducted at a tertiary care hospital, there is a risk of selection bias, as the study population may not fully represent the broader hypertensive population. Second, our analysis was based on complete data from 99,408 out of 168,441 patients. The reliance on electronic health records introduces the potential for information bias due to inaccuracies or incomplete documentation. Third, although we adjusted for several potential confounders, unmeasured variables—such as lifestyle factors (e.g., physical inactivity, diet, smoking, and alcohol consumption) or other interventions—may have influenced the outcomes. Additionally, while the findings may be generalizable to Thai populations in similar healthcare settings, they may not be applicable to other populations due to differences in genetic background, cultural practices, and healthcare systems. Finally, with a median follow-up of 3.7 years, the study may not have captured cases of late-onset NOAF, highlighting the need for longer-term follow-up in future research.

## Conclusions

Older age, male sex, abnormal BMI (both underweight and overweight/obese), and comorbidities are significant risk factors for NOAF in Thai patients with HTN. Statins may have a protective effect, while the role of SGLT2i or GLP-1RA requires further study. Long-term prospective studies are warranted to confirm these findings and guide prevention strategies.

## Data Availability

Data are not publicly available due to sensitivity but can be requested from the corresponding author, Thinnakrit Sasiprapha (Division of Cardiology, Department of Medicine, Faculty of Medicine Ramathibodi Hospital, Mahidol University, 270 Rama VI Road, Ratchathewi, Bangkok, Thailand 10400; email: [insert email]). Data are stored at the CEB Data Warehouse, Ramathibodi Hospital

## Data availability

The data supporting the findings of this study are not publicly available due to their sensitive nature. However, they can be obtained from the corresponding author upon reasonable request. The data are stored in a controlled-access repository at the Department of Clinical Epidemiology and Biostatistics (CEB), Faculty of Medicine Ramathibodi Hospital, Mahidol University, Bangkok, Thailand. Further information regarding the CEB Data Warehouse can be found at https://www.rama.mahidol.ac.th/ceb/CEBdatawarehouse/Overview.

## Author Contributions

V.L. conceptualizations, methodology, data analysis, interpretation, manuscript writing. T.S. methodology, study design, data acquisition, data analysis, interpretation, manuscript revision. H.T. methodology, study design, data acquisition, data analysis, interpretation, manuscript revision. A.P. methodology, data acquisition. S.S. methodology, data acquisition. S.B. methodology, data acquisition. A.T. methodology, data analysis, interpretation, manuscript revision. All authors read and approved the final manuscript.

## Acknowledgments

The study team thanks Dr. Arthur Brown for reviewing the manuscript and providing valuable comments, and CEB Data Warehouse Working Group for their expert advice and support throughout this work.

## Completing interests

The authors had no relevant financial relationships with the medical or pharmaceutical industries.

---

The RECORD statement – checklist of items, extended from the STROBE statement, that should be reported in observational studies using routinely collected health data.

|  | Item No. | STROBE items | Location in manuscript where items are reported | RECORD items | Location in manuscript where items are reported |
| --- | --- | --- | --- | --- | --- |
| Title and abstract |  |  |  |  |  |
|  | 1 | (a) Indicate the study’s design with a commonly used term in the title or the abstract | 1-2 | RECORD 1.1: The type of data used should be specified in the title or abstract. When possible, the name of the databases used should be included. | 2 |
|  |  | (b) Provide in the abstract an informative and balanced summary of what was done and what was found | 2 | RECORD 1.2: If applicable, the geographic region and timeframe within which the study took place should be reported in the title or abstract. | 2 |
|  |  |  |  | RECORD 1.3: If linkage between databases was conducted for the study, this should be clearly stated in the title or abstract. | n/a |
| Introduction |  |  |  |  |  |
| Background rationale | 2 | Explain the scientific background and rationale for the investigation being reported | 2-3 |  |  |
| Objectives | 3 | State specific objectives, including any prespecified hypotheses | 3,6 |  |  |
| Methods |  |  |  |  |  |
| Study Design | 4 | Present key elements of study design early in the paper | 4 |  |  |
| Setting | 5 | Describe the setting, locations, and relevant dates, including periods of recruitment, exposure, follow-up, and data collection | 4 |  |  |
| Participants | 6 | (a) Cohort study - Give the | 4-5 | RECORD 6.1: The methods of study | 4 |
|  |  | eligibility criteria, and the sources and methods of selection of participants. Describe methods of follow-up<br>*Case-control study* - Give the eligibility criteria, and the sources and methods of case ascertainment and control selection. Give the rationale for the choice of cases and controls<br>*Cross-sectional study* - Give the eligibility criteria, and the sources and methods of selection of participants<br>*(b) Cohort study* - For matched studies, give matching criteria and number of exposed and unexposed<br>*Case-control study* - For matched studies, give matching criteria and the number of controls per case | n/a | population selection (such as codes or algorithms used to identify subjects) should be listed in detail. If this is not possible, an explanation should be provided.<br>RECORD 6.2: Any validation studies of the codes or algorithms used to select the population should be referenced. If validation was conducted for this study and not published elsewhere, detailed methods and results should be provided.<br>RECORD 6.3: If the study involved linkage of databases, consider use of a flow diagram or other graphical display to demonstrate the data linkage process, including the number of individuals with linked data at each stage. | n/a<br>8 |
| Variables | 7 | Clearly define all outcomes, exposures, predictors, potential confounders, and effect modifiers.<br>Give diagnostic criteria, if applicable. | 5-6 | RECORD 7.1: A complete list of codes and algorithms used to classify exposures, outcomes, confounders, and effect modifiers should be provided. If these cannot be reported, an explanation should be provided. | 5-6 |
| Data sources/<br>measurement | 8 | For each variable of interest, give sources of data and details of methods of assessment (measurement).<br>Describe comparability of assessment methods if there is more than one group | 5-6 |  |  |
| Bias | 9 | Describe any efforts to address potential sources of bias | 7,16 |  |  |
| Study size | 10 | Explain how the study size was arrived at | 4,8 |  |  |
| Quantitative variables | 11 | Explain how quantitative variables were handled in the analyses. If applicable, describe which groupings were chosen, and why | 5-7 |  |  |
| Statistical methods | 12 | (a) Describe all statistical methods, including those used to control for confounding<br>(b) Describe any methods used to examine subgroups and interactions<br>(c) Explain how missing data were addressed<br>(d) *Cohort study* - If applicable, explain how loss to follow-up was addressed<br>*Case-control study* - If applicable, explain how matching of cases and controls was addressed<br>*Cross-sectional study* - If applicable, describe analytical methods taking account of sampling strategy<br>(e) Describe any sensitivity analyses | 7<br>7<br>16<br>7<br>7 |  |  |
| Data access and cleaning methods |  | .. |  | RECORD 12.1: Authors should describe the extent to which the investigators had access to the database population used to create the study population.<br>RECORD 12.2: Authors should provide information on the data cleaning methods used in the study. | 4,17<br>16 |
| Linkage |  | .. |  | RECORD 12.3: State whether the study included person-level, institutional-level, or other data linkage across two or more databases. The methods of linkage and methods of linkage quality evaluation should be provided. | 6 |
| **Results** |  |  |  |  |  |
| Participants | 13 | (a) Report the numbers of individuals at each stage of the study (*e.g.*, numbers potentially eligible, examined for eligibility, confirmed eligible, included in the study, completing follow-up, and analysed)<br>(b) Give reasons for non-participation at each stage.<br>(c) Consider use of a flow diagram | 8<br>8<br>8 | RECORD 13.1: Describe in detail the selection of the persons included in the study (*i.e.*, study population selection) including filtering based on data quality, data availability and linkage. The selection of included persons can be described in the text and/or by means of the study flow diagram. | 8 |
| Descriptive data | 14 | (a) Give characteristics of study participants (*e.g.*, demographic, clinical, social) and information on exposures and potential confounders<br>(b) Indicate the number of participants with missing data for each variable of interest<br>(c) *Cohort study* – summarise follow-up time (*e.g.*, average and total amount) | 8<br>8<br>9 |  |  |
| Outcome data | 15 | *Cohort study* - Report numbers of outcome events or summary measures over time<br>*Case-control study* - Report numbers in each exposure category, or summary measures of exposure<br>*Cross-sectional study* - Report numbers of outcome events or summary measures | 9 |  |  |
| Main results | 16 | (a) Give unadjusted estimates and, if applicable, confounder-adjusted estimates and their precision (e.g., 95% confidence interval). Make clear which confounders were adjusted for and why they were included<br>(b) Report category boundaries when continuous variables were categorized<br>(c) If relevant, consider translating estimates of relative risk into absolute risk for a meaningful time period | 9-10<br>8-9<br>n/a |  |  |
| Other analyses | 17 | Report other analyses done—e.g., analyses of subgroups and interactions, and sensitivity analyses | 10-11 |  |  |
| **Discussion** |  |  |  |  |  |
| Key results | 18 | Summarise key results with reference to study objectives | 11-12 |  |  |
| Limitations | 19 | Discuss limitations of the study, taking into account sources of potential bias or imprecision. Discuss both direction and magnitude of any potential bias | 16 | RECORD 19.1: Discuss the implications of using data that were not created or collected to answer the specific research question(s). Include discussion of misclassification bias, unmeasured confounding, missing data, and changing eligibility over time, as they pertain to the study being reported. | 16 |
| Interpretation | 20 | Give a cautious overall interpretation of results considering objectives, limitations, multiplicity of analyses, results from similar studies, and other relevant evidence | 12-16 |  |  |
| Generalisability | 21 | Discuss the generalisability (external validity) of the study results | 16 |  |  |
| **Other Information** |  |  |  |  |  |
| Funding | 22 | Give the source of funding and the role of the funders for the present study and, if applicable, for the original study on which the present article is based | 18 |  |  |
| Accessibility of protocol, raw data, and programming code |  | .. |  | RECORD 22.1: Authors should provide information on how to access any supplemental information such as the study protocol, raw data, or programming code. | 17 |

\*Reference: Benchimol EI, Smeeth L, Guttman A, Harron K, Moher D, Petersen I, Sørensen HT, von Elm E, Langan SM, the RECORD Working Committee. The REporting of studies Conducted using Observational Routinely-collected health Data (RECORD) Statement. *PLoS Medicine* 2015; in press.
\*Checklist is protected under Creative Commons Attribution (CC BY) license.

